# Electronic health record enabled track and trace in an urban hospital network: implications for infection prevention and control

**DOI:** 10.1101/2021.03.15.21253584

**Authors:** Li Pi, Paul Expert, Jonathan M Clarke, Elita Jauneikaite, Céire E Costelloe

## Abstract

Healthcare-associated infections represent one of the most significant challenges for modern medicine as they can significantly impact patients’lives. Carbapenemase-producing Enterobacteriaceae (CPE) pose the greatest clinical threat, given the high levels of resistance to carbapenems, which are considered as agents of ‘last resort’ against life-threatening infections. Understanding patterns of CPE infection spreading in hospitals is paramount to design effective infection control protocols to mitigate the presence of CPE in hospitals. We used patient electronic health records from three urban hospitals to: i) track microbiologically confirmed carbapenemase producing *Escherichia coli* (CP-Ec) carriers and ii) trace the patients they shared place and time with until their identification. We show that yearly contact networks in each hospital consistently exhibit a core-periphery structure, highlighting the presence of a core set of wards where most carrier-contact interactions occured before being distributed to peripheral wards. We also identified functional communities of wards from the general patient movement network. The contact networks projected onto the general patient movement community structure showed a comprehensive coverage of the hospital. Our findings highlight that infections such as CP-Ec infections can reach virtually all parts of hospitals through first-level contacts.

## Introduction

Healthcare-associated infections (HCAIs) represent one of the most significant challenges for modern medicine. In Europe, the latest point prevalence report on HCAI detailed incidence estimates between 5.4 to 7.8 per 100 patients^1^, while it is estimated that the overall burden may exceed 15 per 100 patients in resource limited settings^2^. HCAIs can significantly impact patients’ lives, leading to prolonged hospital stays, long-term disabilities and increased risk of mortality. In a recent modelling study, HCAIs were estimated to cause 22,800 deaths and cost the National Health Service (NHS) in England an extra £2.1 billion in 2016/2017^3^. Among healthcare-associated pathogens, Carbapenemase-producing Enterobacteriaceae (CPE), pose the greatest clinical threat, given their high levels of resistance to carbapenems which are considered as agents of ‘last resort’ against life-threatening infections^4^. In the UK, the number of confirmed CPE isolates referred to Public Health England has increased more than 10 fold from 2010 to 2018^5^. Due to this rapid rising trend and the scarcity of alternative antibiotic treatment options, CPE infections have been identified by the World Health Organisation as a critical public health priority, with emphasis on the development of novel therapeutics and improved control measures^6^.

CPE spreads in healthcare settings through contacts between patients, healthcare workers, and contaminated 1 environments^7^. Barring systematic testing, the underlying transmission and colonisation patterns of CPE are difficult to track because of asymptomatic carriage and transmission. This is exacerbated by the pathogens’ ability to persist on surfaces such as patient beds, tables, sink drains, and radiators^8^ which serve as environmental reservoirs to sustain an endemic presence of CPE in clinical environments. Previous studies have focused on investigations and protection measures for patients at high risk of clinically significant CPE infection, with limited resources to examine the specific pathogen infection sources and transmission routes^9^. For example, existing evidence suggests that patients with compromised immune systems, receiving antibiotic therapy or undergoing invasive clinical procedures, are at increased risks of CPE acquisition^7^. To date, most efforts are concentrated on devising efficient testing strategies to protect at risk patient populations^10–12^, while few studies have attempted to explain how patients get infected in a real-world hospital setting by capturing potential transmission dynamics^8,9^.

The widespread adoption of the electronic health records (EHRs) represents a major advancement in healthcare service provision in the 21st century. EHRs provide essential information that can be used to explore spread of CPE and infectious pathogens in general^13^ and in simulation studies^14^. EHRs can be harnessed to record therapeutic procedures, adjustment of diagnosis, microbiology test results as well as time-stamped transfers between wards from admission to discharge. In particular, using microbiology test results and time-stamped transfers between wards, it is possible to determine the spatio-temporal trajectories of confirmed CPE carrier patients, and trace their interaction with other patients —— their contacts.

Networks are commonly used to represent the pairwise interactions of a complex system, and a hospital can naturally be represented as a set of wards linked when patients move from one to another. Analysing the structure of such network will facilitate the understanding of complex patient flows. Networks of patient movements within hospitals are of special interest for HCAIs control because of the great potential for uncovering transmission routes via patients and healthcare workers colonisation and environment contamination related behaviours. While networks models and representations are commonly used in infectious disease epidemiology, both theoretically and practically^15–18^, infection related contact network analysis in hospitals is still an emerging subfield^19,20^ and studies explicitly using a network representation of patient movements focus on local properties of the network^21^, for example, to assign a *C. difficile* susceptibility score to a ward *C. Diff*. or evaluate healthcare delivery performance, such as A&E services^22^, and hospital referrals^23^.

In this study, we explored the practicality of using electronic health records data to identify pathogen transmission patterns using CPE as an example. We used 36 months of EHR data from a multi-hospital urban NHS Trust (a multi-hospital care provider) to investigate potential transmission routes of carbapenemase-producing *Escherichia coli* (CP-Ec) carriers from a network perspective. We describe the population of patients who have contacts with CP-Ec carriers within the NHS Trust, identify the most critical wards involved in potential CP-Ec infection outbreaks, and identify the potential transmission routes for CP-Ec propagation within individual hospitals and across the whole NHS Trust. By considering patient movements within hospitals as a network and monitoring patients’ journeys through this network, fundamental information on CP-Ec transmission characteristics were established. Accounting for the healthcare system complexity, this could serve as an potent way to evaluate the risk of acquiring infections during hospitalisation. In the longer term, precision healthcare, particularly precision infection prevention and control can be proposed based through in-depth exploration of these transmission processes.

## Materials and Methods

### Data resource and study population

#### Hospital and Patient data

We obtained access to de-identified routinely collected individual patient electronic health records from a multi-hospital urban NHS Trust as part of service evaluation(Ref:347). The NHS Trust studied here is one of the largest NHS trusts in England, providing healthcare for more than a million people each year in five hospitals and community services. The Trust comprises five hospital sites, while two of them were excluded from the analysis as they are small-sized specialist maternity and ophthalmology hospitals. The three remaining hospitals accommodate approximately 1,130 acute beds, with 136 beds for intensive care units. There are between 7 to 25 beds in each ward, and 10 wards contain only private rooms. The three hospitals included have different patient populations based on the specialist centres within each hospital. This leads to differences in screening and testing frequency, as screening strategies target primarily clinically at-risk patients^10,11^.

The patient movement dataset contains inpatients information collected over 36 continuous months within the period January 2015 to December 2018, the exact start and end dates were not communicated as part of the Information Commissioner’s Office requirements for data protection. The data include patient demographics, treatment and diagnosis, as well as dates and times for each ward transfer from admission to discharge. In the original dataset, one spell refers to one unique hospitalisation, and each row within a spell corresponds to a change of ward or of main clinical service. We extracted a cleaned dataset in which de-duplicated, timestamped information are stored chronologically. In this process, we removed all spells that presented time continuity inconsistencies or missed essential information such as ward name and transfer time to obtain an unambiguous spatio-temporal representation of patients movement. This leaves the dataset with a total of 219,633 unique patients and 514,753 spells.

A separate microbiology diagnostics dataset for the same population and time span was linked to the patient movement dataset by unique patient ID to identify CPE colonisation status. This microbiology test dataset contains all the microbiology related information, including sample collection date and time, antibiotic sensitivity test, and causative organism.

#### Data availability and ethics statement

De-identified patient data was kept and analysed on a secure server and cannot be made publicly available due to the Information Commissioner’s Office requirements. Access to the datasets used in this paper via a secure environment will be reviewed on request by Imperial College Healthcare NHS Trust. Local institutional ethics oversight body has confirmed no Research Ethics Committee review is required for this project.

#### Study populations

We first defined a general population to illustrate the complex behaviour of flow across the whole Trust and separate hospitals. The general population consisted of all patients admitted as inpatients in the Trust over the observation period, with the exception of the following types of patients: patients admitted in two small-sized specialist maternity and ophthalmology hospitals of the Trust, patients registered in maternity or paediatrics during part their hospitalisation, patients who stayed less than 1 day or longer than 90 days after admission. These inpatients were excluded because they acted as ‘outliers’ of movement networks: either they moved too often in a compressed time period and/or small parts of hospitals, or stayed too long so as to distort the overall traffic pattern. The general population contains data on 55,709 unique patients - out of 219,633 for the whole dataset -, 85,589 unique hospital spells - out of 514,753 for the whole dataset -, including 38,513 with one or more ward transfers, totalling 79,859 ward transfers spanning a total of 160 wards over the 3 hospital sites.

In this study, we focused on *E. Coli* Carbepenem-producing Enterobacteriaceae (CP-Ec) carriers and their contacts, both are subsets of the general inpatient population. The carriers population were identified based on the microbiology diagnostics dataset if *E. Coli* species were detected in any of the tested samples and the antibiotics sensitivity test results showed that the pathogen was resistant to at least one kind of carbapenems (Aztreonam, Ertapenem, Imipenem or Meropenem). The carriers population included both symptomatic and asymptomatic patients.

Based on the movements of the carriers population, we defined the contacts population based on whether they had stayed in the same ward at the same time as a patient from the carriers population up to the carriers positive microbiology sample collection time. This ‘contacts’ population were patients at risk of being colonised with CP-Ec and further spread of CP-Ec, thus, the ward networks connected by the movements of contacts population are the main networks we analysed in this study. Data processing is illustrated in Figure 1 and was conducted using the Pandas 1.1.5 package in Python 3.6.8.

**Figure 1.**
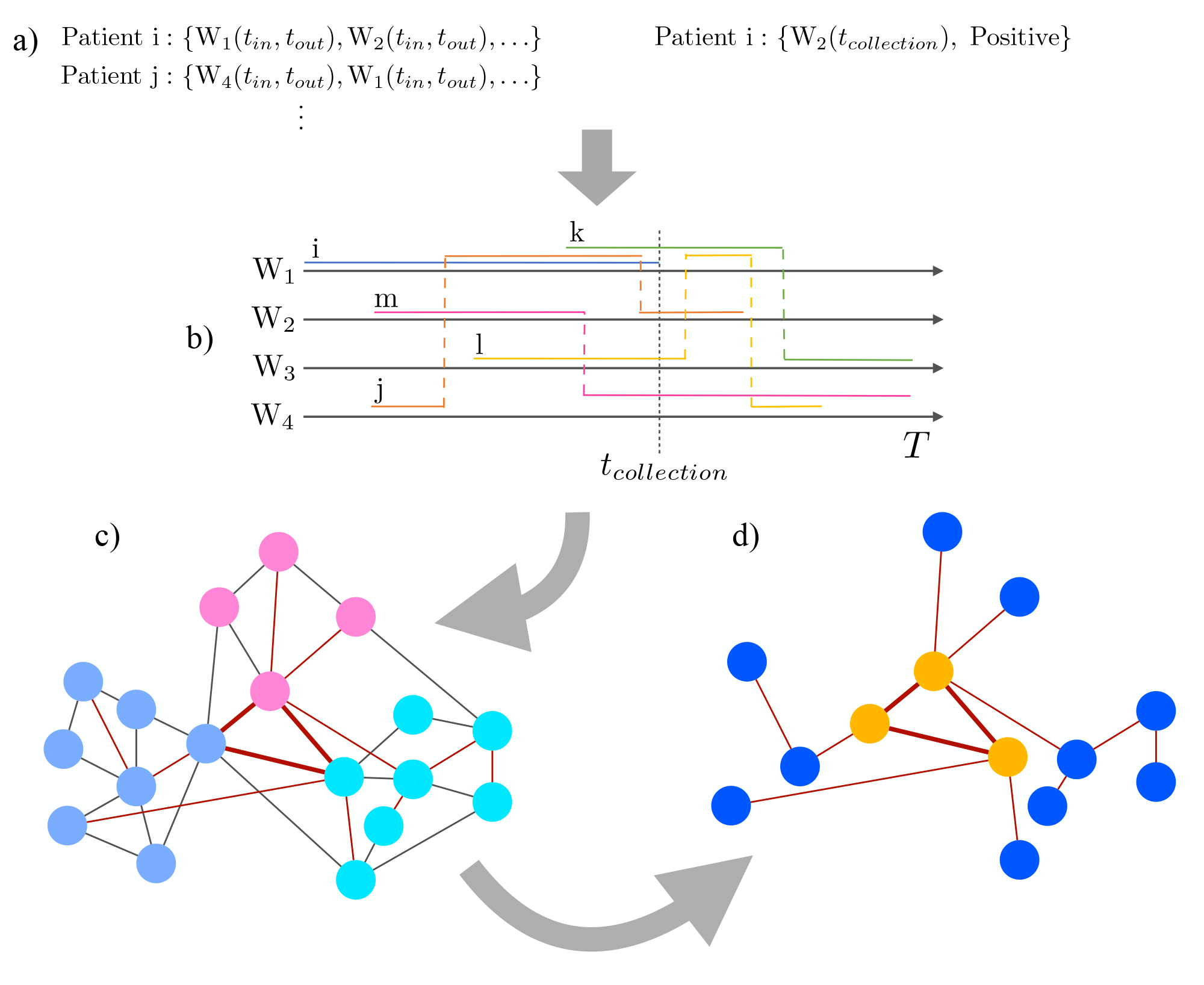
Illustration of the workflow: **a)** Time series of time-stamped ward visits are extracted for all patients, and sample location and collection time is recorded for carrier *i*. **b)** Patients *j,k* are identified as in contacts population because they have been co-located with patient *i* prior to *t*_*collection*_. Patients *m,l* are part of the general population as they never directly share a location with *i*. The trajectory of *i* is ignored post sample collection. **c)** All patient trajectories are collapsed onto the ward network, community structure identified and represented by node colours, contact patient routes are highlighted in red, and ward and community coverage computed. **d)** The contact population network is extracted from the whole population network and the core-periphery structure identified by the node colours, thicker edges correspond to higher transfer count.

### Network construction and analysis

As shown in Figure 1, each spell can be represented as a trajectory: a timestamped sequence of wards visited by the patient. We considered wards as nodes and patient transfers as edges. By iterating through all qualifying spells we constructed undirected, weighted networks of patient movements. The weight of an edge between two nodes represents the frequency of patients having transferred between the two node wards in either direction. We included all wards in the patient movement networks, including procedural wards such as imaging, endoscopy and theatre wards. The ward’s function or clinical speciality was defined by the majority specialised service provided within that ward derived from the Treatment Function Code (TFC) of each spell that qualifies the clinical speciality of the clinician responsible for the patient. For a consecutive period of 12 months, referred to as Year 1, 2 and 3, a contacts network and a background network were constructed for each hospital by aggregating ward transfers of inpatients from the contacts and general inpatient population, respectively. We considered blocks of one year to avoid any seasonal effect.

#### Network metrics

We started by analysing the importance of individual wards in our potential CP-Ec hospital transmission networks, which could be quantified by network metrics measuring how ‘central’ or ‘important’ a node is within a network. For the 9 contacts networks (three hospital sites and three years) and 9 background networks, we computed several classical nodal graph metrics: *strength, closeness, betweenness, eigenvector centrality, clustering* and *eccentricity*. Each metric provides a different measure of the centrality, or importance, of a node in a network.

In a weighted network, the *strength* of a node is defined as the sum of the weights of its incident edges. In the context of this paper, the strength of a node corresponds to the number of patient transfer to/from the ward. *Closeness* and *eccentricity* both measure how close a node is to others and are based on the notion of shortest path between two nodes. The shortest path between two nodes is defined as the path with the largest sum of weights, i.e. the higher the weight connecting two nodes, the closer they are. *Closeness* is the average value of the shortest path length from a node to every other node, while *eccentricity* is calculated as the reciprocal of the ‘longest’ shortest path length from a node to all the others. *Betweenness* is defined by the proportion of times a node sits on the shortest path length of two other nodes and quantify how important one node is in controlling information flow in the network assuming information follows shortest paths. By contrast, *eighenvector centrality* quantify the importance of a node by taking into account the importance of the nodes it is connected to. For example, a ward connected with many other well-connected wards would have higher eighenvector centrality than a ward with same number of less well-connected neighbours. *Clustering* refers to the local clustering coefficient, and measures how the strength of the connectivity between the neighbours of a node. The detailed calculations and interpretations of these metrics can be found in Newman’s textbook^24^.

We used Spearman’s rank-order correlation to measure the correlation between the ward centrality metrics, the number of contacts between the carriers and contacts population up to the time of the carriers identification in the ward, and the number of carriers identified in the ward. Wards with equal metric value are given equal rank.

#### Mesoscopic structures

We also evaluated the modular, or community, structure of wards by measuring 2 types of node block structures: the community structure and the core-periphery structure. The community structures were measured on the background networks to identify groups of wards based on the volume of patients transferred. In this context, communities were effectively coherent functional subsets of wards. The core-periphery structures were measured on the contacts networks to assess the extent to which a set of nodes tends to act as interaction hubs. Each spell is associated with a Treatment Function Code (TFC) that reflect the clinical speciality of the clinician responsible for the patient. A ward is assigned the TFC corresponding to the most common TFC of the patients visiting it. The same method is applied to assign a type of function to each cluster.

Community structure represents coherent sub-networks and is loosely defined as finding sets of nodes that are more connected among themselves than with the rest of the network. When present, the adjacency matrix of the network is block diagonal when nodes are ordered by community assignment. Communities were found by optimising modularity^25^ using the Clauset-Newman-Moore algorithm^26^. Assigning a community to a node allows the definition of another node based metric: the *participation coefficient*^27^. This metric measures the extent to which a node is connected to nodes outside its own community and can act as an inter community hub.

The core-periphery structure, in its simplest form, is a generalisation of the star graph structure and can be represented in block form as 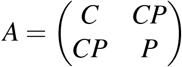, with the sum of the weights in the block *C* larger than the sum of the weights in the block *CP* and much larger than in the block *P* (||*C*|| ≤ ||*CP*|| « ||*P*||)^28^ The core nodes are densely connected together and form the analogue to the central node in the star graph. The core and peripheral nodes are also strongly connected, while the nodes in the periphery are comparatively less densely connected, completing the analogy with the star graph. The nodes of a network possessing a core-periphery structure can thus be assigned a core or periphery tag, and edges classed into three categories based on the tags of their end nodes: core, core-periphery and periphery. In the present work, we are interested in understanding the structure of wards with high traffic and connectivity with respect to wards with a lower diversity of traffic/connectivity. To uncover the node core-periphery structure, we used an agglomerative algorithm. It is initialised by selecting the heaviest edge in the network and assigning both its end nodes to the core. Two steps are then iterated until the weight of the core edges account for the majority of the total network weight: i) the set of edges connected to a core node, excluding already used edges, is created ii) the heaviest edge of the set created in i) is selected, if it connects to a node not in the core set, that node is added to it. The pertinence of the core-periphery structure is assessed by the ratios of the core weights to the periphery weights. The similarity of the core-periphery assignment between two years was measured using the Hamming distance for the ward present in all years. The Hamming distance measures the proportion of core and periphery ward assignment changes between two years. For a comprehensive discussion of core-periphery structure, see Kojaku and Masuda^28^.

Analysis were performed using Python 3.6.8: Package Pandas were used for numerical calculation and the Networkx 2.5 and igraph 0.8.3 packages were used for network metrics calculation. Graphs were produced using Gephi (version 0.9.2). Codes for core-periphery characterisation are available at xxxc

## Results

### Study populations

Our study population included 55,709 individual patients, 85,589 admissions, 79,859 transfers spanning a total of 160 wards from an urban Trust comprising 3 hospitals collected over 36 continuous months during the period January 2015 to December 2018. The patient characteristics of interest were described in Tables SI 1, SI 2 and SI 3 for each hospital site. For the carriers population, defined as patients with at least one positive microbiology sample and included both symptomatic and asymptomatic patients, the statistics on ward transfers only included transfers up to the collection time of the first positive microbiology sample. The number of CP-Ec carriers we identified in each hospital were between 10 and 92 per year, with only (12,12,10) patients classified as carriers in Hospital C. Carriers have a median ward transfer of 2 - except for two occurrences, see Table SI 1, during their hospitalisations in each year, though their contacted patients accounted for up to 2% to 17% of total inpatients admitted each year across hospitals. The clinical characteristics of patients are different across three hospitals, as reflected by the distribution of Treatment Function Code, a field that characterise the speciality of the consultant responsible for a patient.

### Networks metrics

9 contacts networks and 9 background networks were constructed using movements of contacts population and general population, respectively. The sets of wards for each hospital network is determined by the wards visited by the patients forming the general population. Maternity and paediatrics patients were filtered out according to their registered treatment function code and thus mostly exclude wards associated with these specialities, but in rare instances a patient that is not seen by maternity/paediatrics consultant might still be transferred to a maternity/paediatrics ward, leading some of these wards to be included in the networks and the ward set to vary marginally across the 3 years. Another driver of change in ward sets is ward decommissioning. For each undirected, weighted network, wards were ranked based on node importance measured by metrics introduced in the Method section, as well as by the number CP-Ec carriers identified in a ward and the number of contacts between the carrier and contact population up to the time of the carriers identification. As shown in Figure 2, we found that there was no strong consistent Spearman-rank correlations between these ward metrics, which suggests that the nodes properties may not be the optimal indicators of wards’ CP-Ec risk. However, among the 6 metrics we measured, strength and eigenvector centrality were relatively more likely to be positively correlated with ward CP-Ec carriage risk in both contacts networks and background networks. This can be explained because strength directly measures the ward traffic by summing the total number of transfers from/to the corresponding ward, and eigenvector centrality compounds this information to account for the strength of the wards it exchanges patients with.

**Figure 2.**
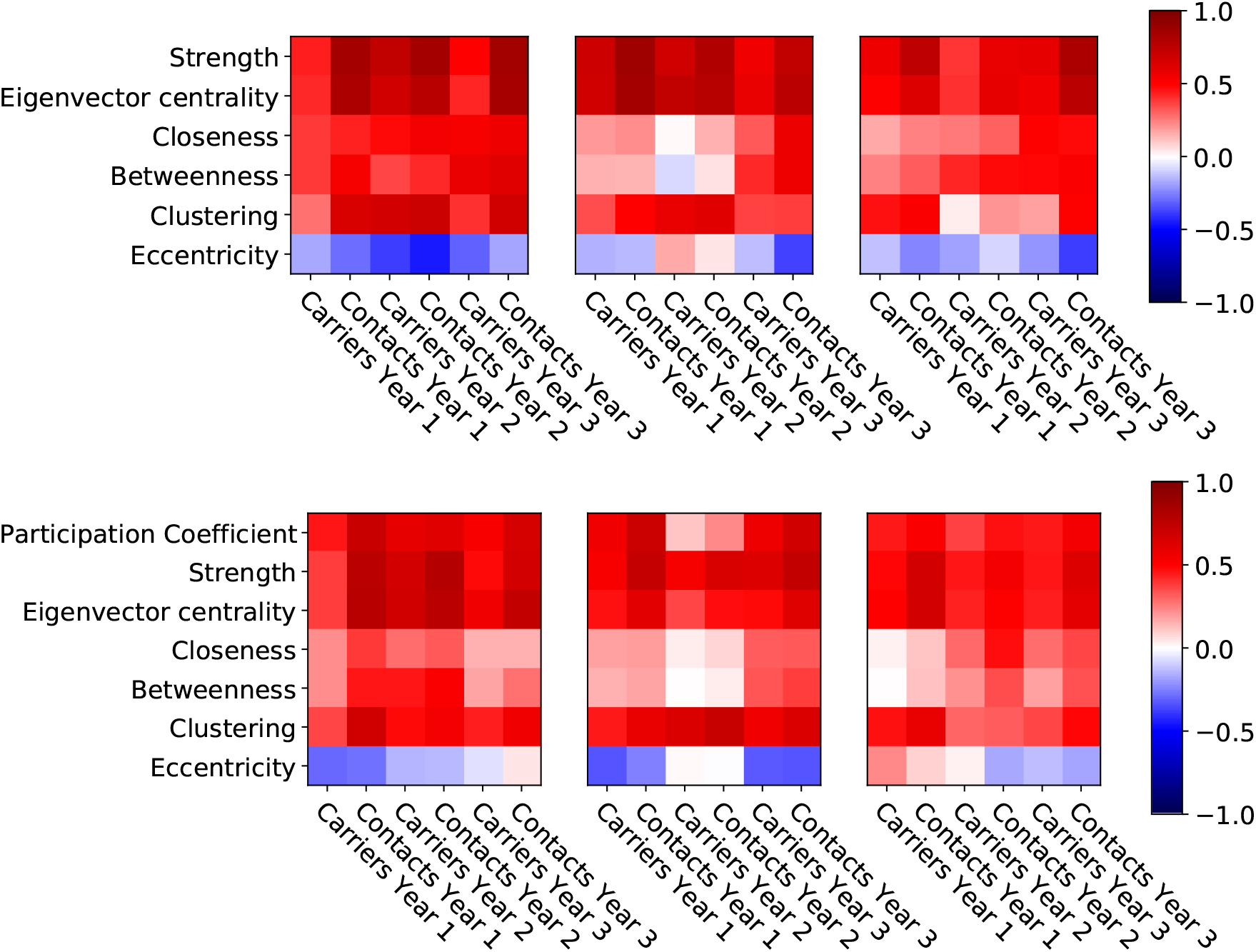
Heatmaps of the Spearman rank-correlations between the ward network metrics and the number of carriers identified in a ward, and the number of contacts between carriers and contacts in a ward up to the carriers identification time in the contacts network (Top) and the background network (Bottom). Columns are by year, and hospitals A,B and C from left to right.

### Background networks and hospital community structure

Communities are groups of wards that exchange significantly more patients among themselves and were obtained by optimising modularity of the background networks of each hospital and year. Communities are consistently present over time, with values ranging from 0.30 to 0.50, see Table 1. As expected, the community structure closely followed ward specialities, a reflection of established clinical pathways, with wards from one or more speciality grouped together. The right column of Figure 3 shows the wards community assignment by colours for each hospital in Year 2. The ward labels represent the dominant treatment function code of the patients visiting the ward and community labels can be obtained from the top three treatment speciality of the patients of the wards they comprise, but communities can contain wards from other specialities.

**Table 1.** Modularity values for each hospital and each year, with the number of communities in parenthesis.

|  | year 1 | year 2 | year 3 |
| --- | --- | --- | --- |
| Hospital A | 0.41 (4) | 0.41 (5) | 0.46 (6) |
| Hospital B | 0.52 (4) | 0.50 (4) | 0.47 (6) |
| Hospital C | 0.47 (6) | 0.42 (6) | 0.30 (7) |

**Figure 3.**
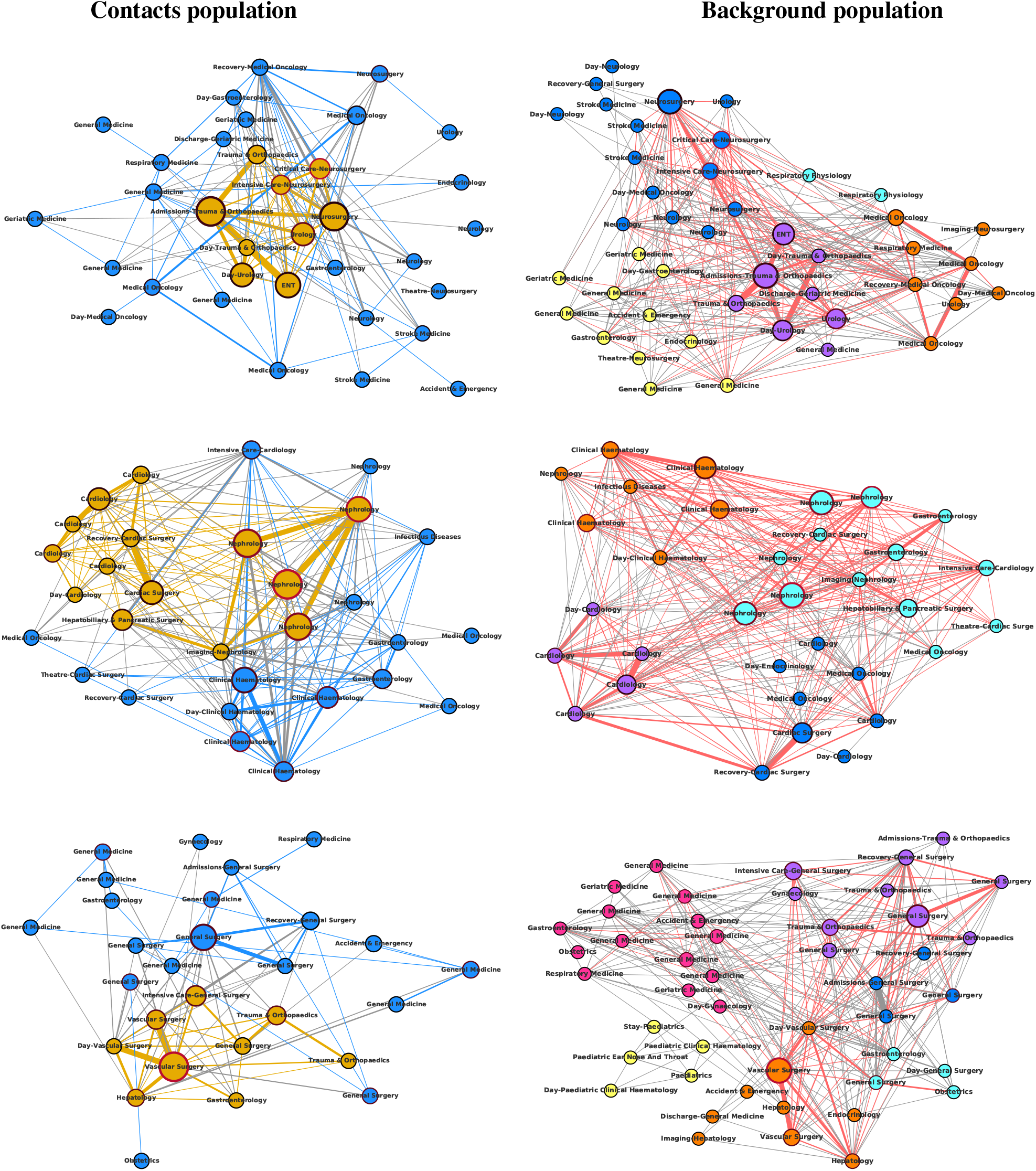
(Left column) patient movement of contacts population aggregated over Year 2. Core nodes are in yellow, peripheral nodes in blue. The size of the nodes is proportional to the number of interactions - same time and place - between carriers prior to collection time and contacts. The redness of the border of the node is proportional to the number of carriers detected in the node. Edges between core nodes: yellow, edges between core and peripheral nodes in grey, edges between peripheral nodes in blue. (Right column) patient movement of background population aggregated over Year 2. The node colour represents community assignment. The size of the nodes is proportional to the number of interaction - same time and place - between carriers prior to collection time and contacts. Edge colour is grey if no patient from the contact population moved between a pair of nodes, red if at least one such patient moved between a pair of nodes. The red edges are the same as in the edges of in the left column.

We noted that although community structure was consistently present, wards exact community assignment did vary in time as shown in Figure SI 1. These variations in assignment were marginal and were mostly due to the fact that some wards accommodate patients from a wide range of specialities and modularity being a hard partitioning method. For example, there are several private wards in Hospital B; this private wards community was only detected in Year 3 while they merged with another ‘Oncology & Endocrinology’ community in Year 1 and 2 when their patients received more similar services as Oncology & Endocrinology patients. Other functional wards, such as imaging, admission, discharge wards, were also naturally ‘floated’ across different communities. Patient flow management intervention could also explain why certain sub-speciality communities emerges, e.g. the ‘Respiratory Medicine’ in hospital A Year 2 that then persist in Year 3. The variation in the sets of wards across time is a result the wards visited by the general population, see Networks Metrics section.

### Contacts networks and core-periphery structure

The core-periphery structure in the contacts population movement networks was revealed using the agglomerative algorithm introduced in. We observed a robust core-periphery structure, which was stable across the 3 years and 3 hospitals. Of particular interest were the Core-Periphery ratios that shows the pertinence of the core-periphery classification for Hospital A (6.54,4.56,5.01), B (3.28,2.73,3.80) and C (1.85,2.34,2.64). The detailed core-periphery structure statistics were shown in Table SI 4. These ratios were stable over time, while the ratios for Hospital A were higher compared to that in Hospital B and C, which indicates that the core-periphery structure observed in Hospital A is more noticeable. The Hamming distance between the core periphery assignments between the wards present in the contacts network across the three years, see Table SI 5, further confirmed the stability of the core periphery structure.

As an example, we focus the discussion in this section on Year 2. The node colour of left column of Figure 3 shows the node core-periphery assignment for each hospital during Year 2. The speciality composition of wards in each Core and Periphery clusters were shown in Figure SI 2. We observed that they were highly heterogeneous, in contrast to the communities which contain mainly wards of similar speciality, confirming that the core-periphery structure captureed a different type of mesoscopic organisation to the community structure. The main specialities of core wards were consistent with the top specialities of wards visited by carriers (see Table SI 1), while also contained other wards which were not traditionally considered as wards at high risks of CP-Ec clinical infections. We noted that not all wards were present in the core-periphery networks, as they were built upon the wards visited by carriers and contacts, which might vary over time and not cover the whole hospital. We observed that as with the community assignment variations, these were negligible and the core-periphery structure was significantly present as highlighted above.

### Hospital coverage

We measured hospital coverage by the carriers and contacts using the proportion of wards and the proportion of communities reached by each population. Carriers visited at least 39%, 63% and 30% of wards in hospital A, B and C respectively, and these minimal percentages went up to 64%, 80% and 44% in the contacts population. When considering community coverage, the minimum was 71% across all hospitals and reached 100% in some cases, both for the carriers and contact populations(for example, Year 1 in Hospital A). The complete list of figures was reported in Table 2. We also presented in Figure SI 3 the statistics for the number of wards and communities visited by carriers and contacts during time at risk, defined as time spent in hospitals for contacts and time to sample collection for carriers. In Figure 3, red edges correspond to ward transfer used at least once by a patient from the contact population and gives a visual representation of the community coverage by this population.

**Table 2.**
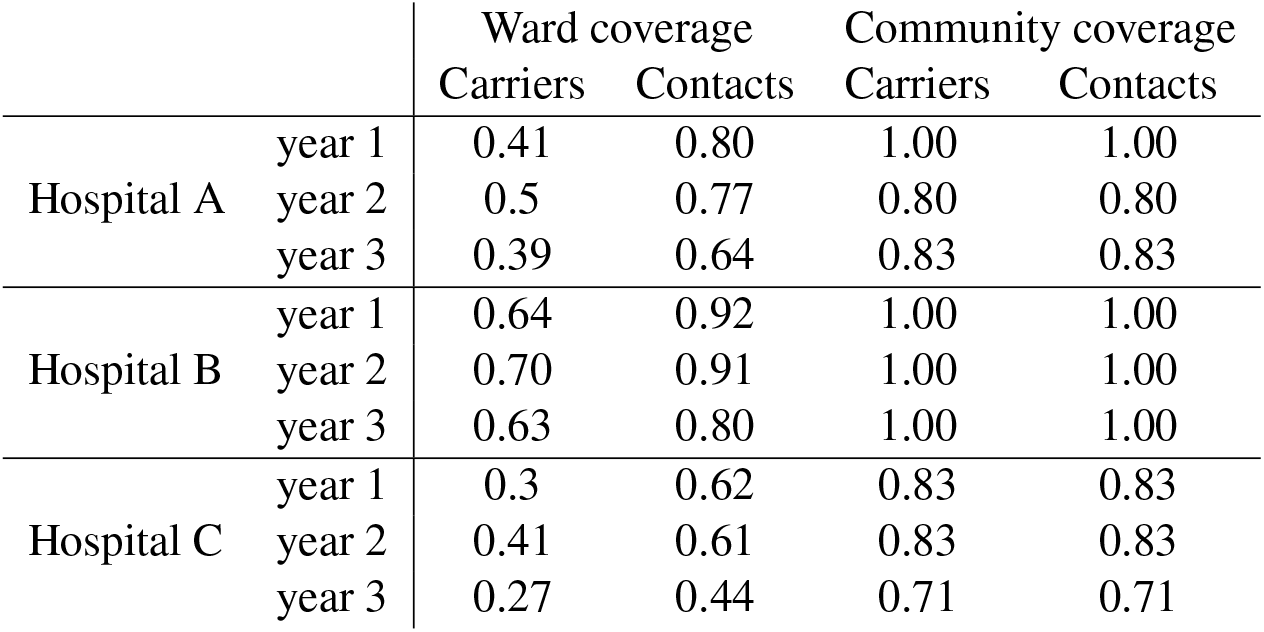
Coverage per hospital/year of the ward by cases/at-risk as well as communities reach by cases/at-risk. We see that communities reached is constant in the two population, but the coverage of wards is significantly increased by the at-risk population.

## Discussion

### Summary of results

HCAIs are a known threat to patient safety in hospitals and their control and prevention are an integral part of patient care^29^. CPE is an endemic problem within hospital systems and there are no effective decolonization options^30^. CPEs are often resistant to many first-line antibiotics, which limits treatment options and contributes to high morbidity and mortality rates related to infection^31^. Due to these complexities it is important to reduce the risk of vulnerable CPE-naive patients acquiring CPE during their hospitalisation. Identification of areas within a hospital with high patient interactions will allow identification of specific settings for targeted infection prevention and intervention. However monitoring of the entire patient population for potential infection is unfeasible. An alternative is to monitor how confirmed carriers move within the hospital network, and how they interact with infection-free or asymptomatic patients that came in contact with carriers^32^. Studying these movement patterns in a ‘track and trace’ style approach may provide valuable insights into more effective patient management and infection control.

Our proposed track and trace approach explicitly consider hospitals as complex systems to reveal that there are common patient movement patterns that are stable in time, and across hospitals studied, in a large urban NHS Trust. A core-periphery structure is systematically present in the contacts population networks of all hospitals, indicating a strong presence of “patient highways” between core wards where most interactions between carriers and contacts occur. Carriers and contacts patients are then redistributed to the peripheral wards, associated with varied patient populations and located across well-defined functional clusters of wards, with the potential to spread organisms further. Interestingly, a high prevalence of carriers is not necessarily associated with core wards, supporting the mixing and disseminating picture. Additionally, wards with a relatively higher number of carriers correspond to specialities where screening or systematic testing of patients occurs: nephrology, cardiology^10,11^, and at least partially explain the difference in observed prevalence.

The potential for the transmission of a pathogen through the hospital and across specialities is confirmed by the analysis of the community structure of the whole patient population and its coverage by the contact population with the presence of peripheral wards in most communities, see Table 2. Community structure, or clustering, is a common feature of complex systems^33^ that usually constrains epidemic and diffusive dynamics^34–36^ that in the case of patient movement and hospital wards highlight functional clusters comprising wards that commonly share patients. The communities found in the three hospitals indeed are thematically homogeneous, see Figure SI 1. However, while wards within the same community tend to share more patients than with wards in other communities, cross community patient transfer does occur, and in particular, the contacts population not only reach most communities, but also a large number of wards in each community, see Figure3. Furthermore, as the three hospitals are part of the same Trust, we observed that both carriers (before confirmation of CP-Ec) and contacts are transferred between hospitals, see SI 4, increasing the risk of cross hospital contamination and colonisation by CP-Ec. This shows that pathogens can reach any functional group wards using only primary contacts with carriers. Indeed, the results presented in our study may represent a ‘best case scenario’ as we have not considered secondary and higher order contacts, contacts with healthcare workers, or delayed transmission, which would only further extend the coverage of pathogens.

### Limitations

We acknowledge several limitations of our study. We have taken a pragmatic approach to define interactions, based on contacts spending time in the same ward at the same time as a carrier. We are also limited by the spatial resolution of measurements. Pilot studies using Radiofrequency ID(RFID) tag like approaches to observe patient interactions^37^ and patient-healthcare workers-administrative staff^38^ would partially alleviate this limitation, but this is not easily scalable and could raise privacy considerations. Simulation studies exploring finer interaction patterns and calibrated using real data could lead to more precise estimates of potential spread of pathogens.

Another limitation is variation in the sampling rate of the hospital population for CPE infection. While we do not know how many tests are ordered for CP-Ec, there is a clear disparity in positive CP-Ec patient between hospitals, e.g. hospital B in our study which has a between 65 and 92 confirmed carriers and hospital C which has between 10 and 12 confirmed carriers SI 1. This might be due to the patient population in hospital B being more at risk of developing infection and therefore prevention testing being more systematic. Under the assumption that there is no difference between hospital population in the risk of being colonised by CP-Ec, this implies that asymptomatic carriers may be moving freely, undetected within more general hospitals such as hospital C. This is detrimental to building a clear picture of potential CP-Ec colonisation in hospital, particularly since the three hospital we analysed here are strongly interconnected: pathogens can easily migrate far from wards treating at-risk patients.

Finally, we have considered CPE in pertaining to *Escherichia coli* only in this study. CPE colonisation Enterobacterales are a large family including species such as *Klebsiella spp* and *Enterobacter spp* therefore we are not presenting a complete picture of movements across the plethora of CPE carriers and contacts.

## Conclusions and recommendations

We draw several conclusions from our study. First, our results show the importance of strong infection control protocols, given the potential reach and spread of CPE. Despite this reach, the number of CPE carriers in our dataset were low, testament to the adherence to robust existing infection prevention and control policy, but perhaps also reflecting the low sampling rate among asymptomatic carriers. Second, it makes clear that the studied hospitals possess a core-periphery structure distributing patients across functional clusters, limiting the protective effect community structure has on infection spreading^35^. While different pathogens may spread by different routes, e.g. airborne or surface contamination, and the timing of detection of asymptomatic carriers and symptomatic patients could give rise to pathogen-specific patterns, they are likely to share general common properties such as core-periphery structure and hospital coverage particularly if higher order contact tracing are considered.

Finally, in the longer term, precision healthcare could incorporate information on patient movement patterns to enhance screening activity. Currently, in the Trust, enhanced screening has been operational periodically to screen patients who are admitted to specialities considered to be high risk (adult and paediatric ICU, adult and paediatric haematology, and renal), who live overseas or who have had an overnight stay in a UK hospital in the past 12 months, and patient contacts of known CPE carriers who could not be isolated^29^. An extension to include random screening, including lower risk patient groups could be incorporated into infection prevention and control practice for a better understanding of prevalence in hospital in services that are not the target of systematic screening.

This study brings a novel application of electronic health record information, utilising network modelling methods to study patient movement patterns in a multi-site hospital network. Using a track and trace approach the results unveiled the movement patterns of carriers and their interaction with contacts. This movement traffic reveals a clear core-periphery structure which covers the entire hospital network, with implications for the ease of spread of infection through these patient traffic routes. The methods demonstrated here leverage data collected routinely and therefore could be readily incorporated into routine hospital surveillance operations. The implications of infection prevention control measures, for example ward closures could be studied in terms of their impact on infection transmission. Furthermore, the results suggest that routine screening programmes could be adapted to be random and with wider coverage. As CPE becomes endemic in hospital systems worldwide, surveillance systems need to adapt. Utilising EHR data, readily available due to the digitalisation of healthcare systems, combined with network analysis represents and additional tool in the arsenal of defence against antimicrobial resistant pathogens such as CPE.

## Data Availability

We obtained access to de-identified routinely collected individual patient electronic
health records from a multi-hospital urban NHS Trust as part of service evaluation(Ref:347). De-identified patient data was kept and analysed on a secure server and cannot be made publicly available due to the Information Commissioner's Office requirements. Access to the datasets used in this paper via a secure environment will be reviewed on request by Imperial College Healthcare
NHS Trust. Local institutional ethics oversight body has confirmed no Research Ethics Committee review is required for this project.

## Acknowledgements

This research was funded by the NIHR Imperial Biomedical Research Centre (NIHR-BRC-P68711). The research was conducted using National Institute of Health Research Health Informatics Collaborative data resources. Data management (and analytical support) was provided by the Big Data and Analytical Unit (BDAU) at the Institute of Global Health Innovation (IGHI). The authors are also thankful to Dr C. Coughlan for their clinical advice. CC is supported by a personal NIHR Career Development Fellowship (grant number NIHR-2016-090-015). PE is supported by the NIHR Imperial BRC (grant number NIHR-BRC-P68711), PE and JC acknowledges support from EPSRC grant EP/N014529/1 supporting the EPSRC Centre for Mathematics of Precision Healthcare. JC acknowledges support from the Wellcome Trust (215938/Z/19/Z). EJ is a Rosetrees/Stoneygrate 2017 Imperial College Research Fellow, funded by Rosetrees Trust and the Stoneygate Trust. Imperial College London is grateful for the support from the North West London NIHR Applied Research Collaboration.The views expressed in this publication are those of the authors and not necessarily those of the NIHR or the Department of Health and Social Care.

## Author contributions statement

Conceptualisation: CC, PE

Formal Analysis: LP, PE Funding acquisition:

CC Investigation: LP, PE

Methodology: LP, PE

Clinical expertise: JC

Microbiology: LP, EJ

Software: LP, PE

Visualization: LP, PE

Writing – original draft: LP

Writing – review & editing: All authors

## Additional information

### Ethics

Ethics for this study was granted service evaluation approval through Imperial College London NHS Trust (Ref:347). The datasets were fully anonymised and compliant with Information Commissioner’s Office requirements for de-identification. Imperial College Joint Research Office is the dedicated ethics oversight body that has confirmed the study is exempt from Research Ethics Committee review.

The authors declare that they have no competing interests.

## Supplementary Information

### Tables

**Table SI 1.** Carriers population statistics for all 3 hospitals and 3 years. Interquartile ranges for the: Ward transfers, Length of stay; standard deviation for age.

|  | Year 1 | Year 2 | Year 3 |
| --- | --- | --- | --- |
| Hospital A |  |  |  |
| Length of stays | 9.46 (IQR:6.03-27.27) | 9.30 (IQR:3.19-19.91) | 9.72 (IQR:6.41-19.28) |
| Ward transfers | 2 (IQR:2-3) | 2 (IQR:1-2) | 1 (IQR:1-3) |
| Age | 60.95 (SD:15.65) | 62.2 (SD:12.63) | 61.3 (SD:15.23) |
| Gender (F/M) | 5/16 | 25/16 | 11/19 |
| TFC 1 | Neurosurgery (6) | Neurosurgery (10) | Neurosurgery (10) |
| TFC 2 | Urology (4) | Urology (9) | Urology (4) |
| TFC 3 | Trauma & Orthopedics (2) | Medial Oncology (6) | Trauma & Orthopedics (4) |
| #Spells | 21 | 41 | 30 |
| #Patients | 21 | 39 | 29 |
| Hospital B |  |  |  |
| Length of stay | 16.58 (IQR:7.90-30.07) | 13.40 (IQR:6.09-27.47) | 9.72 (IQR:5.95-22.05) |
| Ward transfers | 2 (IQR:1-3) | 2 (IQR:1-3) | 2 (IQR:1-3) |
| Age | 62.44 (SD:13.59) | 61.86 (SD:15.79) | 63.08 (SD:15.64) |
| Gender (F/M) | 27/63 | 44/59 | 31/44 |
| TFC 1 | Nephrology (43) | Nephrology (50) | Nephrology (21) |
| TFC 2 | Clinical Haematology (20) | Clinical Haematology (27) | Clinical Haematology (19) |
| TFC 3 | Cardiology (8) | Cardiology (11) | Cardiology (13) |
| #Spells | 90 | 103 | 75 |
| #Patients | 79 | 92 | 65 |
| Hospital C |  |  |  |
| Length of stay | 20.09 (IQR:11.88-25.19) | 16.12 (IQR:7.95-26.03) | 12.78 (IQR:4.78-19.07) |
| Ward transfers | 2 (IQR:1-3) | 1 (IQR:1-2) | 2 (IQR:1-2.25) |
| Age | 60.92 (SD:18.62) | 65.75 (SD:18.14) | 67.50 (SD:6.65) |
| Gender (F/M) | 3/9 | 3/9 | 5/5 |
| TFC 1 | Vascular Surgery (3) | Vascular Surgery (3) | General Surgery (3) |
| TFC 2 | General Surgery (2) | General Surgery (2) | Trauma & Orthopedics (3) |
| TFC 3 | Respiratory Medicine (2) | General Medicine (2) | Vascular Surgery (1) |
| #Spells | 12 | 12 | 10 |
| #Patients | 12 | 12 | 10 |

**Table SI 2.** Contacts population statistics for all 3 hospitals and 3 years. Interquartile ranges for the: Ward transfers, Length of stay; standard deviation for age.

|  | Year 1 | Year 2 | Year 3 |
| --- | --- | --- | --- |
| Hospital A |  |  |  |
| Length of stay | 5.38 (IQR:2.46-11.22) | 4.31 (IQR:1.83-9.27) | 4.34 (IQR:2.22-9.52) |
| Ward transfers | 2 (IQR:2-3) | 2 (IQR:2-3) | 2 (IQR:2-3) |
| Age | 60.28 (SD:15.65) | 59.26 (SD:15.52) | 59.36 (SD:15.82) |
| Gender (F/M) | 226/192 | 438/374 | 256/217 |
| TFC 1 | Neurosurgery (95) | Neurosurgery (190) | Neurosurgery (133) |
| TFC 2 | Trauma & Orthopedics (91) | Urology (156) | Trauma & Orthopedics (119) |
| TFC 3 | ENT (52) | Trauma & Orthopedics (115) | ENT (60) |
| #Spells | 418 | 812 | 473 |
| #Patients | 414 | 794 | 167 |
| Hospital B |  |  |  |
| Length of stay | 8.75 (IQR:4.32-16.22) | 8.17 (IQR:4.18-16.08) | 8.00 (IQR:3.60-15.40) |
| Ward transfers | 3 (IQR:2-4) | 3 (IQR:2-4) | 3 (IQR:2-4) |
| Age | 59.09 (SD:15.21) | 57.87 (SD:16.34) | 58.52 (SD:17.31) |
| Gender (F/M) | 540/843 | 655/1025 | 528/800 |
| TFC 1 | Nephrology (644) | Nephrology (744) | Nephrology (429) |
| TFC 2 | Cardiology (226) | Clinical Haematology (374) | Cardiology (313) |
| TFC 3 | Clinical Haematology (190) | Cardiology (255) | Clinical Haematology (308) |
| #Spells | 1383 | 1680 | 1328 |
| #Patients | 1246 | 1418 | 1157 |
| Hospital C |  |  |  |
| Length of stay | 11.08 (IQR:5.01-20.09) | 10.74 (4.85-22.12) | 15.06 (6.10-20.29) |
| Ward transfers | 3 (IQR:2-4) | 3 (IQR:2-4) | 2 (IQR:2-4) |
| Age | 60.72 (SD:17.67) | 60.90 (SD:19.05) | 61.01 (SD:18.07) |
| Gender (F/M) | 74/117 | 32/88 | 40/56 |
| TFC 1 | Vascular surgery (63) | Vascular Surgery (40) | General Surgery (32) |
| TFC 2 | General Surgery (40) | General Surgery (27) | Colorectal Surgery (16) |
| TFC 3 | Trauma & Orthopedics (20) | Colorectal Surgery (18) | Vascular Surgery (10) |
| #Spells | 191 | 120 | 96 |
| #Patients | 189 | 120 | 96 |

**Table SI 3.** Background population statistics for all 3 hospitals and 3 years. Interquartile ranges for the: Ward transfers, Length of stay; standard deviation for age.

|  | Year 1 | Year 2 | Year 3 |
| --- | --- | --- | --- |
| **Hospital A** |  |  |  |
| Length of stay | 1.30 (IQR:0.48-3.99) | 1.35 (IQR:0.48-4.10) | 1.32 (IQR:0.47-3.74) |
| Ward transfers | 1 (IQR:1-2) | 1 (IQR:1-2) | 1 (IQR:1-2) |
| Age | 55.35 (SD:16.91) | 55.64 (SD:16.77) | 55.71 (SD:16.85) |
| Gender (F/M) | 5703/5092 | 5842/5254 | 5515/5030 |
| TFC 1 | Medical Oncology (2237) | Medical Oncology (1780) | Medical Oncology (1999) |
| TFC 2 | Urology (1497) | Urology (1581) | Urology (1364) |
| TFC 3 | Neurosurgery (1269) | Trauma & Orthopedics (1484) | Trauma & Orthopedics (1334) |
| #Spells | 10799 | 11096 | 10545 |
| #Patients | 8269 | 8731 | 8152 |
| **Hospital B** |  |  |  |
| Length of stay | 2.10 (IQR:0.97-6.95) | 1.91 (IQR:0.65-6.17) | 1.86 (IQR:0.46-6.28) |
| Average ward transfers | 2 (IQR:1-3) | 1 (IQR:1-3) | 1 (IQR:1-3) |
| Age | 59.51 (SD:16.21) | 58.29 (SD:16.55) | 57.8 (SD:16.66) |
| Gender (F/M) | 4209/5655 | 5327/6976 | 5460/6877 |
| TFC 1 | Cardiology (3366) | Cardiology (3639) | Cardiology (3569) |
| TFC 2 | Nephrology (2245) | Nephrology (2704) | Nephrology (2729) |
| TFC 3 | Hepatobiliary & Pancreatic Surgery (892) | Cinical Haematology (1716) | Clinical Haematology (1973) |
| #Spells | 9865 | 12124 | 12337 |
| #Patients | 7393 | 8416 | 8424 |
| **Hospital C** |  |  |  |
| Length of stay | 1.21 (IQR:0.27-3.62) | 1.10 (IQR:0.25-3.26) | 0.52 (IQR:0.20-2.43) |
| Ward transfers | 1 (IQR:1-2) | 1 (IQR:1-2) | 1 (IQR:1-2) |
| Age | 52.65 (SD:20.02) | 53.02 (SD:19.73) | 53.12 (SD:19.23) |
| Gender (F/M) | 2583/3421 | 2801/3442 | 2988/3341 |
| TFC 1 | General Surgery (1876) | General Surgery (1861) | General Surgery (2276) |
| TFC 2 | Vascular Surgery (639) | Endocrinology (776) | Endocrinology (834) |
| TFC 3 | Trauma & Orthopedics (624) | Trauma & Orthopedics (688) | Trauma & Orthopedics (657) |
| #Spells | 6005 | 6243 | 6329 |
| #Patients | 5073 | 5291 | 5258 |

### Figures

**Figure SI 1.**
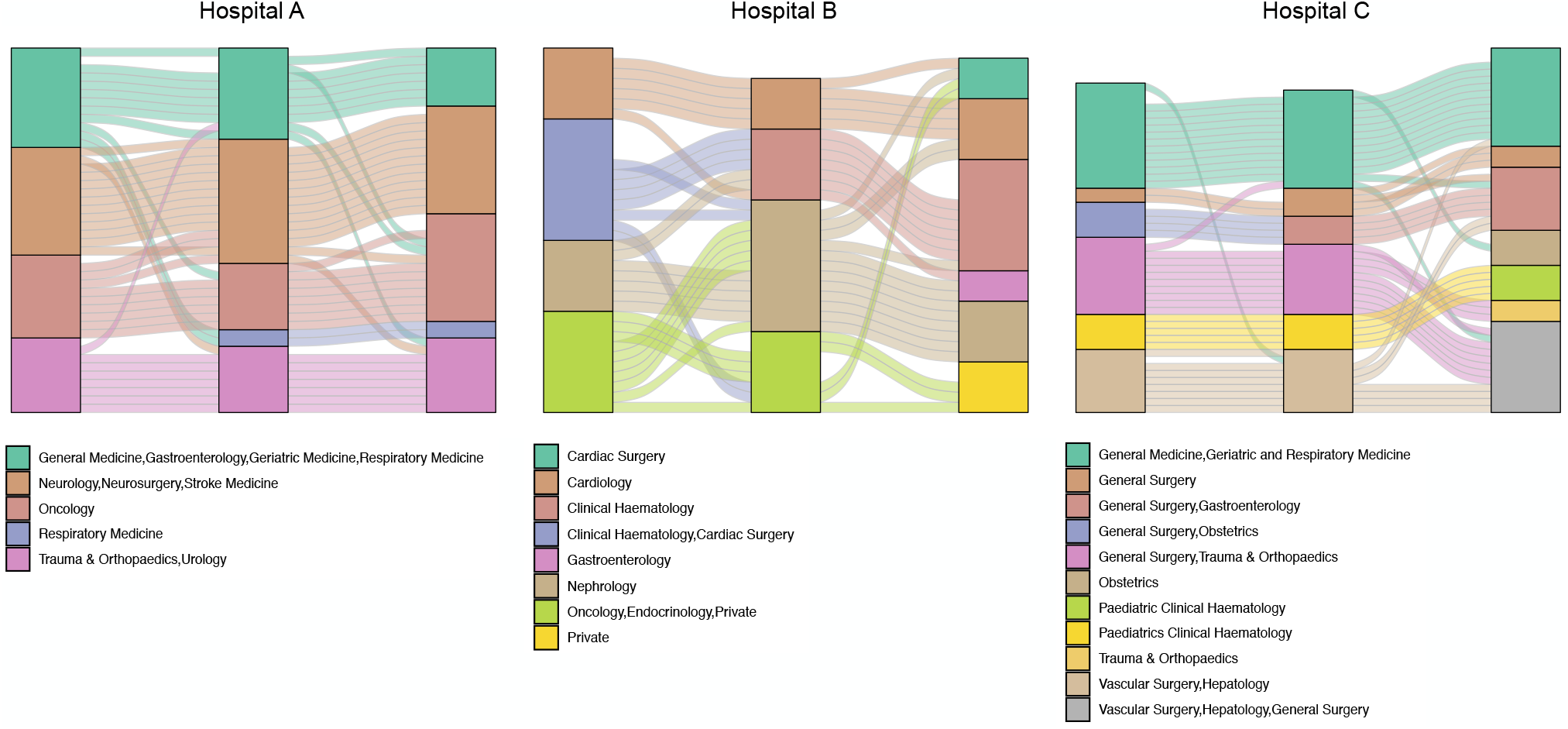
Evolution of the community structure over the three years in each hospital. Community labels represent the top three specialities of the wards comprising a community.

**Figure SI 2.**
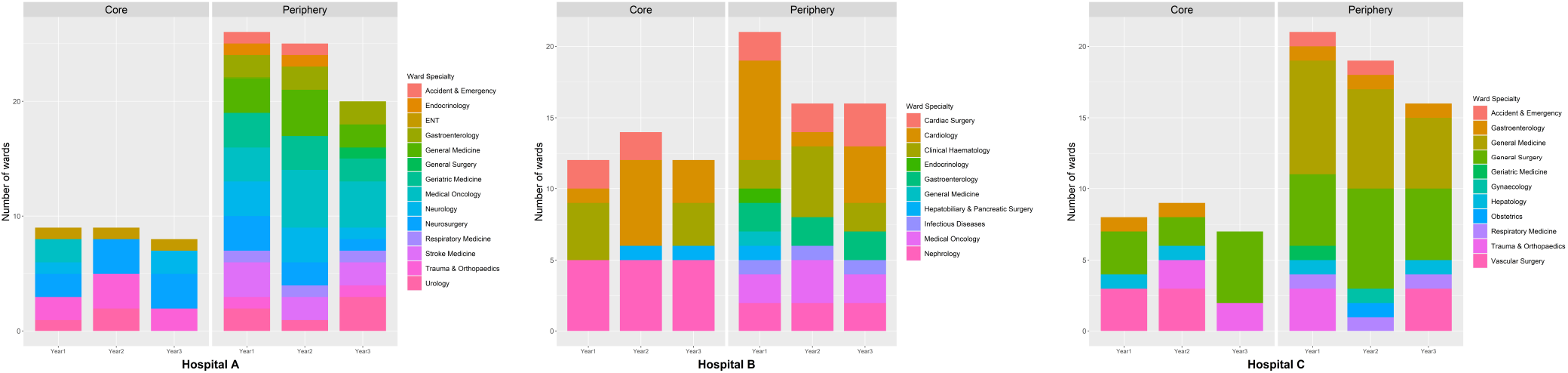
Core and periphery ward assignments for three hospitals in three years

**Figure SI 3.**
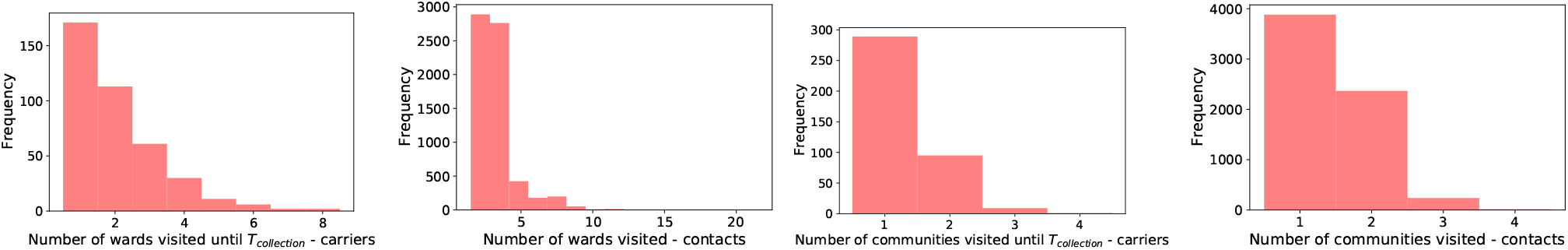
Histograms from left to right: Number of wards visited by carriers prior to *T*_*coll*_, number of wards visited by contacts, number of communities visited by carriers prior to *T*_*coll*_, number of communities visited by contacts

**Table SI 4.**
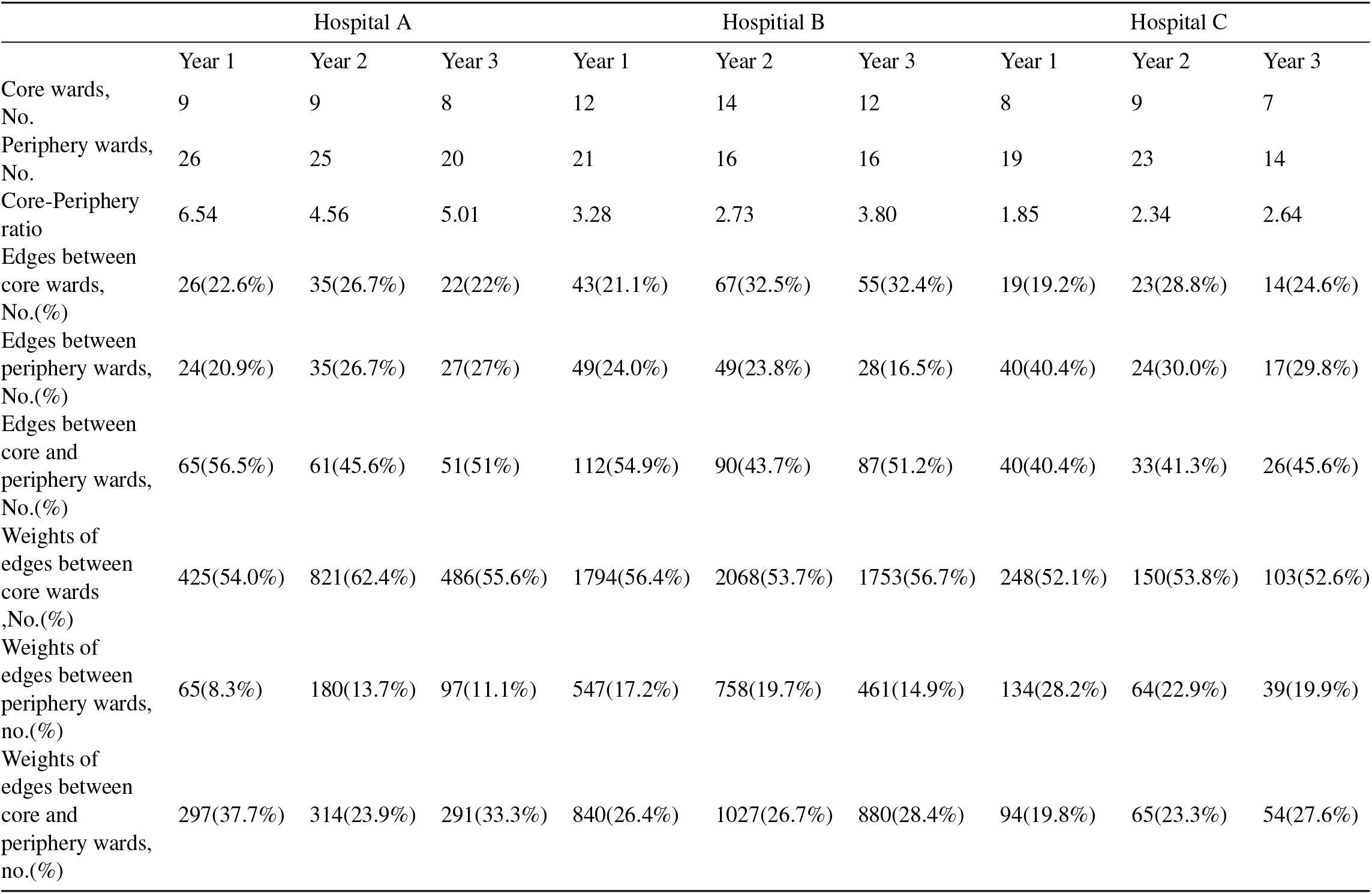
Core-periphery structure statistics for contacts networks

|  | Hospital A |  |  | Hospital B |  |  | Hospital C |  |  |
| --- | --- | --- | --- | --- | --- | --- | --- | --- | --- |
|  | Year 1 | Year 2 | Year 3 | Year 1 | Year 2 | Year 3 | Year 1 | Year 2 | Year 3 |
| Core wards, No. | 9 | 9 | 8 | 12 | 14 | 12 | 8 | 9 | 7 |
| Periphery wards, No. | 26 | 25 | 20 | 21 | 16 | 16 | 19 | 23 | 14 |
| Core-Periphery ratio | 6.54 | 4.56 | 5.01 | 3.28 | 2.73 | 3.80 | 1.85 | 2.34 | 2.64 |
| Edges between core wards, No.(%) | 26(22.6%) | 35(26.7%) | 22(22%) | 43(21.1%) | 67(32.5%) | 55(32.4%) | 19(19.2%) | 23(28.8%) | 14(24.6%) |
| Edges between periphery wards, No.(%) | 24(20.9%) | 35(26.7%) | 27(27%) | 49(24.0%) | 49(23.8%) | 28(16.5%) | 40(40.4%) | 24(30.0%) | 17(29.8%) |
| Edges between core and periphery wards, No.(%) | 65(56.5%) | 61(45.6%) | 51(51%) | 112(54.9%) | 90(43.7%) | 87(51.2%) | 40(40.4%) | 33(41.3%) | 26(45.6%) |
| Weights of edges between core wards ,No.(%) | 425(54.0%) | 821(62.4%) | 486(55.6%) | 1794(56.4%) | 2068(53.7%) | 1753(56.7%) | 248(52.1%) | 150(53.8%) | 103(52.6%) |
| Weights of edges between periphery wards, no.(%) | 65(8.3%) | 180(13.7%) | 97(11.1%) | 547(17.2%) | 758(19.7%) | 461(14.9%) | 134(28.2%) | 64(22.9%) | 39(19.9%) |
| Weights of edges between core and periphery wards, no.(%) | 297(37.7%) | 314(23.9%) | 291(33.3%) | 840(26.4%) | 1027(26.7%) | 880(28.4%) | 94(19.8%) | 65(23.3%) | 54(27.6%) |

**Table SI 5.** Hamming distance matrices between the wards constituting the core for each pair of years and the three hospitals.

**Table SI 4.** Core-periphery structure statistics for contacts networks
|  | Hospital A |  |  |  | Hospital B |  |  |  | Hospital C |  |  |
| --- | --- | --- | --- | --- | --- | --- | --- | --- | --- | --- | --- |
|  | year 1 | year 2 | year 3 |  | year 1 | year 2 | year 3 |  | year 1 | year 2 | year 3 |
| year 1 | 0.00 | 0.24 | 0.20 | year 1 | 0.00 | 0.42 | 0.31 | year 1 | 0.00 | 0.15 | 0.55 |
| year 2 | 0.24 | 0.00 | 0.20 | year 2 | 0.42 | 0.00 | 0.35 | year 2 | 0.15 | 0.00 | 0.40 |
| year 3 | 0.20 | 0.20 | 0.00 | year 3 | 0.31 | 0.35 | 0.00 | year 3 | 0.55 | 0.40 | 0.00 |

**Figure SI 4.**
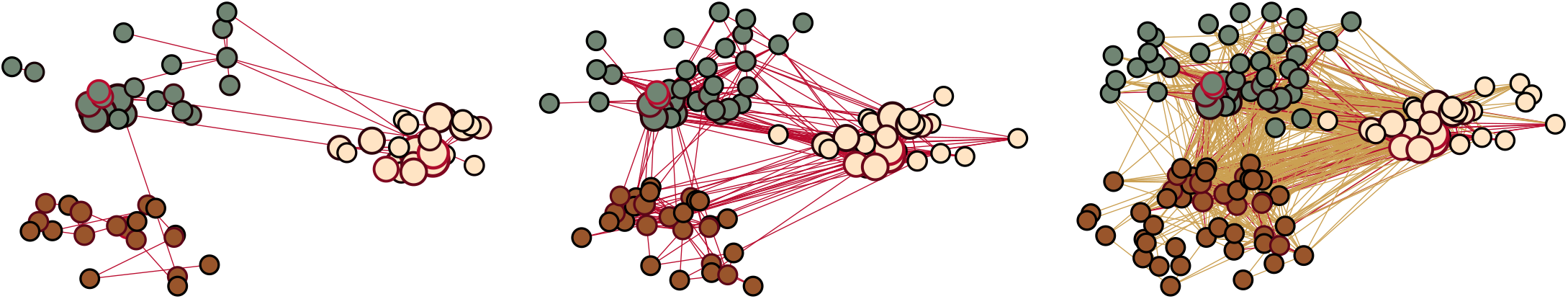
Left to right: Carriers movements, contacts movements, background population movements. The colour of a node represents hospital. Node border colour: proportional to the number of cases.

